# Socioeconomic Status and Depression – A Systematic Review

**DOI:** 10.1101/2023.12.04.23299380

**Authors:** Anders Jespersen, Rebecca Madden, Heather C. Whalley, Rebecca Reynolds, Stephen M. Lawrie, Andrew M. McIntosh, Matthew Iveson

## Abstract

**Objective:** Low socioeconomic status (SES) has been associated with an increased risk of depression and psychiatric disorders in general. In this systematic review and meta-analysis, we aim to provide an estimate of the risk of clinical depression associated with low SES across cultures, age groups and study designs. Finally, we tested whether associations between SES and depression differed by the income of the country in which the study was conducted.

**Methods:** A literature search across five databases returned 7,943 studies. Title, abstract and full text screening resulted in 162 included studies of which 122 were meta-analysed, 22 were included in a cross-sectional narrative review and 19 studies were included in a longitudinal narrative review. Meta-analyses were divided into risk estimates for composite SES, income, education, and employment. Sensitivity analyses based on differences in economic situation in the country of study origin were performed to investigate a possible source of between study heterogeneity.

**Results:** Low SES was associated with an increased risk of depression across all measures of SES. Low income was associated with the highest odds ratio for depression (OR = 1.96, 95% CI = 1.53-2.52). Sensitivity analyses revealed no significant differences in between-study heterogeneity or risk of depression between high- and low-income economy groups.

**Conclusions:** Comparable risks of depression across economy groups suggest that income relative to your peers, rather than absolute income, is a risk factor for depression. Preventative measures and possible policy interventions are discussed.

**Strengths and limitations of this study:**

- This systematic review provides the largest and most comprehensive review and meta-analysis of the association between socioeconomic status and depression.
- The included studies span a broad range of ages, cultures, and country economies, allowing for better generalisation of the results.
- The inclusion of component parts of socioeconomic status (income, education, and employment) in the meta-analysis allows for comparisons of the different risk estimates.
- The broad inclusion criteria are likely to allow for increased between-study heterogeneity.
- Due to the observational nature of the studies included it is difficult to make conclusions on the direction of causality between socioeconomic status and depression. However, the inclusion of a longitudinal narrative review may give an indication of a direction of causality.

## Introduction

Depression is a leading cause of disability world-wide, affecting over 322 million people (WHO 2017). Depression is associated with an elevated mortality risk (Gilman et al., 2017) due, in part, to an increased risk of suicide in depressed people (Bachmann 2018), and to a number of common life-shortening comorbidities (Walker et al 2018.). While there is a significant genetic contribution to the risk of developing depression (Sullivan et al 2000) there is evidence that environmental risk factors, amongst them poverty, deprivation, and low socioeconomic status, are likely to confer an increased risk of depression.

Socioeconomic status (SES) refers to a composite level of income, education and occupation, where lower SES reflects overall material and social deprivation. Lower SES has previously been associated with an increased risk of developing depression (Lorant et al., 2003) and is one of the strongest predictors of overall poor mental and physical health (Everson et al., 2002). Low SES is also a risk factor for many of the chronic diseases associated with or comorbid with depression (Hosseinpoor et al., 2012; Robbins et al., 2005). This relationship may be bidirectional as chronic disease can lead to loss of income, unemployment, or interrupted education.

Many studies of psychiatric disorders in general and depression in particular control for SES when investigating the effects of other risk factors of interest. However, this approach minimises the contribution of poverty itself, which may have larger effects on depression than many of the factors being studied. For example, a recent meta-umbrella systematic review (Arango et al. 2022) on modifiable risk factors common to major psychiatric disorders failed to include analysis of any socioeconomic measures.

While a growing body of research links low SES with poor health, there is a global trend for increasing life expectancy, rising levels of education and fewer people living in poverty (Roser & Ortiz-Ospina, 2016; Roser et al., 2013). However, this trend is not uniform across all countries (World Bank Group 2020; Marmot 2020).

Given the persistent nature of relationships between low SES and health outcomes, such as increased risk of depression, there is a clear need to quantify the strength of this association. It is also unclear whether the relationship of depression with SES depends upon an individual’s absolute socioeconomic position, or on their position relative to that of other individuals in the same country. Previous research on the association between SES and depression have shown evidence of an association in specific cultures and contexts (Richardson et al. 2015), certain age groups (Reiss et al 2013), and sections of society (Lund et al 2010).

In the present review, we systematically reviewed the existing literature measuring the effects of low SES on risk of depression. We provide an synthesis of studies from countries spanning the spectrum of income economies. We sought to test whether differences in SES were associated with the absolute level of poverty in the nation from which participants were recruited, or if differences in SES were consistent across income settings and therefore indicative of an association with relative rather than absolute social position.

## Methods

### Inclusion and exclusion criteria

Peer-reviewed literature and published PhD theses that examined the association between SES and clinically relevant depression were included. PhD theses were included to mitigate publication bias.

Studies involving participants with clinical depression were included if they used any recognised structured diagnostic criteria, whether self-reported, obtained through data linkage, or diagnosed using a diagnostic clinical interview or questionnaire with a clear clinical cut-off. Studies focusing on symptoms and comparisons between subgroups of depression were excluded. Studies with an unclear division between cases and controls i.e. comparing sub groups of depressions, and those with insufficient results to be meta-analysed were also excluded if study authors were not able to provide this information. There were no restrictions on the age, sex, cultural heritage or comorbid diseases of the participants in the present review.

Studies that grouped depression with other psychiatric disorders (often anxiety or schizophrenia) in the “case” group and compared it to a “control” group without both the psychiatric disorders present in the “cases” group were excluded. However, there were no restrictions on comorbidities being included in the present systematic review. As a result, studies with participants with more than one psychiatric disorder or health problem were included, as long as the only difference between the cases-group and controls-group was the presence or absence of depression.

### Search strategy

Literature searches were performed in the following databases: PsycInfo, EMBASE, Medline, Web of Science, ASSIA. The databases were chosen to cover as broad a range of medical and sociological sources as possible.

In four out of five databases searches were conducted using subject headings. The following search template was used in PsycInfo, EMBASE. Medline and ASSIA: Depression AND (Socioeconomic status OR Income OR Occupation OR Education).

Web of Science does not facilitate subject heading searches. Instead, a keyword search was conducted using the same key words that were used in the subject heading search (Depression ADJACENT (Socioeconomic status OR Income OR Occupation OR Education)) ensuring the keywords were within 5 words of each other using the “adjacent” search function.

In all databases, filters were applied to exclude reviews, conference abstracts and animal research and to limit the search to English language papers. There were no restrictions on publication date or country of origin of the search results. Studies on perinatal depression, depression subtypes and depressive symptoms were not filtered out. This was done to minimise automatic exclusion of studies meeting inclusion criteria. Instead, studies investigating perinatal depression, depression subtypes and depressive symptoms were excluded during the two screening processes.

The last date for all searches was 15^th^ of March 2019.

A protocol for this systematic review and meta-analysis was published on the international prospective register of systematic reviews (PROSPERO). The protocol can be found under the ID: CRD42019129509.The protocol was followed except for a risk of bias assessment. To date there is no universally recognised screening tool for risk of bias for observational studies (Bero et al., 2018) and the most popular tools for observational research have been found to be inconsistent (Losilla et al. 2018). The Newcastle-Ottawa Scale for case control and cohort studies (mentioned in the protocol) has been argued to include invalid items, producing highly arbitrary results (Stang 2010). As a consequence the authors of this review decided not to conduct a formal risk of bias assessment.

### Data collection

Abstracts and titles were initially screened by one author (AJ). Studies screened that met inclusion criteria but could not be found through online databases were requested through the University of Edinburgh’s interlibrary loan services. Assessing interrater reliability is important to ensure that the screening process does not miss any eligible studies and that the screening method is replicable across different assessors. Selection bias can be introduced in the screening processes if an assessor repeatedly includes or excludes studies based on factors other than those listed in the inclusion and exclusion criteria. To assess interrater reliability and selection bias a second author (RM) screened a 10% subsample (N = 795) of abstracts and titles. There was an 89.7% agreement on the outcome of the screening by the two authors. 521 studies were screened against their full text.

Data collection was done alongside full text screening, and a sample of the data collection sheet can be found in Additional file 1.

### Analysis

For the meta-analyses the studies were divided into four groups according to how they measured SES: dichotomous, ordinal with three groups, continuous, and longitudinal. The dichotomous study group was made up of all the studies reporting data in a binary/dichotomous format and all the studies reporting results in an ordinal format with 4 socioeconomic groups. The latter type of study was dichotomised by merging the top two socioeconomic groups together (high) and the bottom two socioeconomic groups together (low). Dichotomous, studies were meta-analysed separately according to the type of SES indicator used: a composite SES measure, income, education, and employment (unemployment). As a result of meta-analysing each SES indicator separately, some studies appeared in several meta-analyses, because they reported results for more than one indicator.

A pooled OR and 95% CI was calculated for each SES indicator using the package ‘meta’ (Balduzzi et al., 2019) in R version 4.0.4. The effects were calculated using a random effects model and the between studies variation was calculated using the Sidik-Jonkman estimator (IntHout et al., 2014). RRs and ORs were meta-analysed together as they are statistically comparable when the prevalence is low.

Data files and code can be accessed at: https://osf.io/dqnp5/?view_only=ca92b1486ab842b4bbf8c20f9ea4e4c2.

A narrative approach was adopted for papers that could not be meta-analysed due to the design of the study or insufficient reporting of results. Similarly, all longitudinal papers were included in a longitudinal narrative review due to a combination of low number of studies in each of the SES groups and variable follow-up time.

Significant heterogeneity between studies was anticipated given the large number of studies included and the range of countries, age groups and diagnostic tools reported. Between study heterogeneity was explored using Cochrane’s Q and quantified by an I^2^ statistic. Both measures were calculated using the ‘meta’ package in R. To explore possible reasons for between-study effect size heterogeneity, sensitivity analyses based on the economic situation in the country of origin were performed. The World Bank country and lending group classifications were used to divide countries into low- and high-income economy subgroups. A sensitivity analysis was carried out for each measure-specific meta-analysis on dichotomised data to compare ORs and CIs and heterogeneity measures between high- and low-income economy subgroups. We hypothesised that estimates of depression risk associated with SES indicators would be similar across low- to high-income economies, consistent with the importance of relative position within a country, rather than absolute differences.

## Results

### Study inclusion

The literature search returned 8,312 studies, 369 of which were duplications. During title and abstract screening of the 7,943 remaining studies, 7,422 were excluded. The remaining 521 studies were full text screened, leading to another 359 papers being excluded. A breakdown of the reason for exclusions can be seen in Additional file 2.

As a result, 162 studies were included in the systematic review and meta-analysis. Figure 1 shows a flow-chart with the number of studies excluded at each step and the number of included studies in each of the meta-analyses.

**Figure 1:**
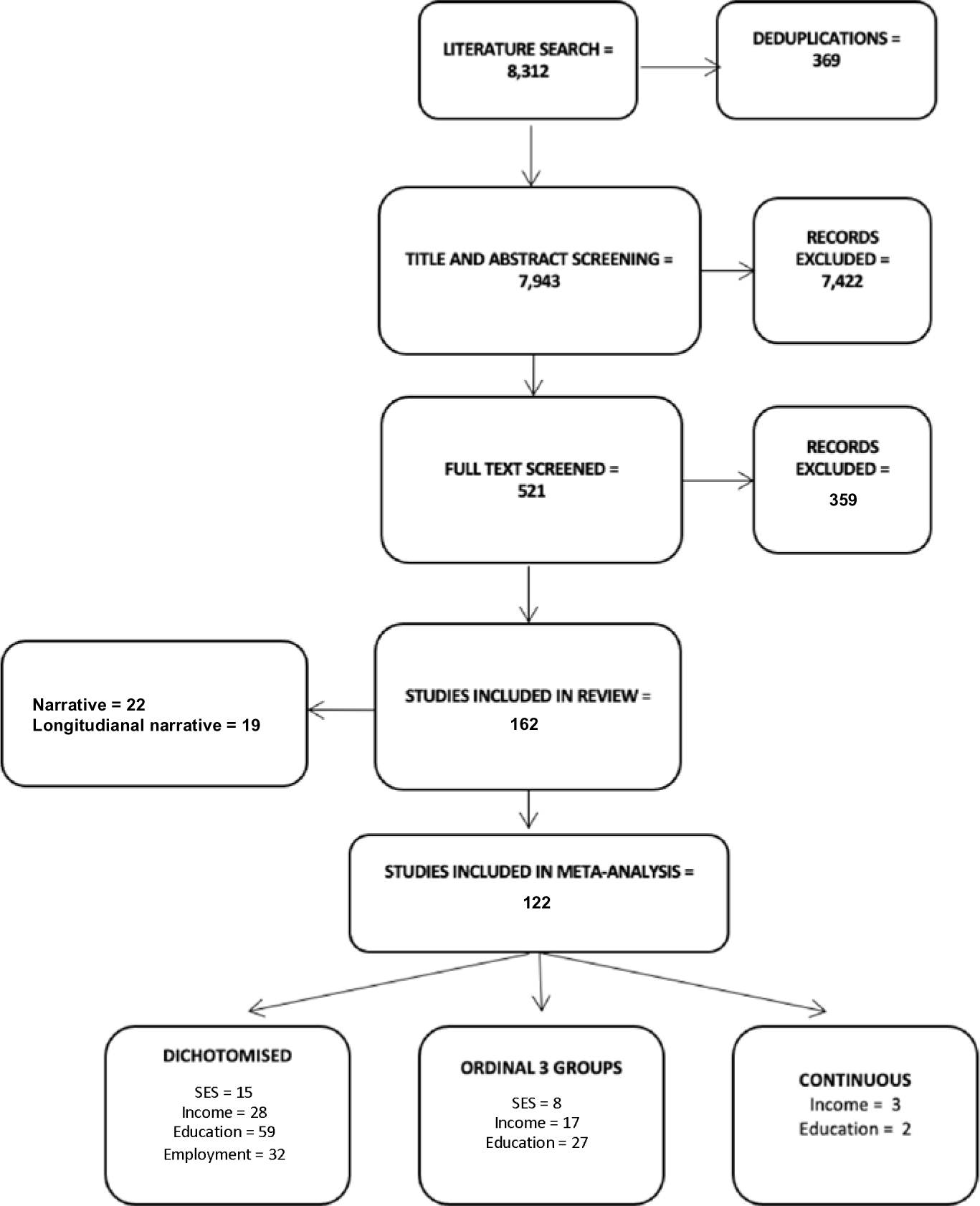
Flowchart of the number of included and excluded studies. Studies in the “Dichotomised” bin reported data in 2 groups (e.g. high vs. low income), studies in the “ordinal 3 groups” bin reported data in 3 groups (e.g. low, medium, and high educational attainment), and studies in the “continuous” bin reported continuous results (e.g. median income for depressed people).

### Missing data and contact with authors

A total of 17 studies reported insufficient data (only worded or graphed results, inconsistencies in reported numbers, no reporting of cut-off, etc.) to include the study in one of the meta-analyses or one of the narrative reviews. Authors of 15 out of the 17 studies were contacted for data or clarification. Author contact details for 1 study could not be found and another had since deceased. Out of the 15 authors that were contacted, 7 replied, and 1 provided supplementary data enabling the study to be included in the meta-analysis.

### Study population

The total number of participants in the 162 studies was 329,005 of which 48,158 presented with depression. The resulting overall prevalence is 14.6%. The participants were recruited from 47 countries across 6 continents.

Of the studies included in the meta-analysis, 63 were from high income countries, 30 from higher middle-income economies, 11 from lower middle-income economies and finally 10 from low-income economies.

In 56 of the studies reporting statistics on their participants age, participants were over 18 and had a mean age below 50. Participants had a mean age of over 50 in 31 of the studies. Children were included as participants in 10 of the studies.

Finally, of the studies included in the meta-analysis 72 of them recruited participants from a community setting while 39 of them recruited from a clinical setting.

### Dichotomised meta-analyses

#### Socioeconomic status

Results from the 15 studies included in the dichotomised meta-analysis of the association between composite SES measures and depression can be seen in Figure 2. The pooled OR for composite SES and depression was 1.70 (95% CI = 1.32-2.20), representing a 70% increase in the odds of depression among individuals of low SES compared to high SES. A sensitivity analysis showed no significant differences in the estimates of risk between high- and low-income economy subgroups (df = 1, p = 0.67)

**Figure 2:**
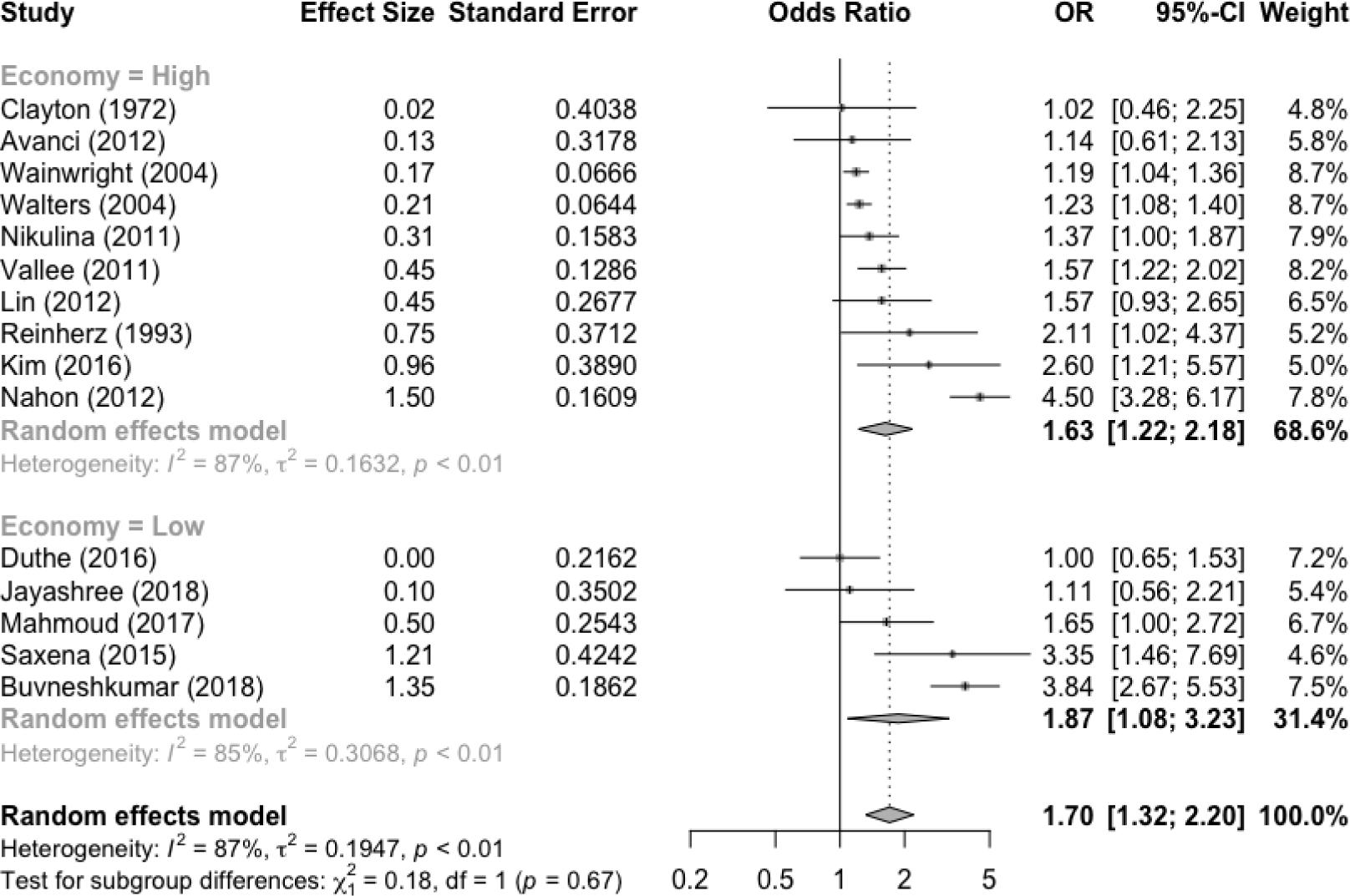
Forrest plot of ORs and CIs of depression risk in people with low SES, from dichotomised data. (e.g. high vs. low SES). Results are shown for each individual study, a pooled estimate for high- and low-income economy subgroups, and pooled estimate for all included studies.

The between-study heterogeneity was estimated at I^2^ = 87% (with a variance of *τ*^2^ 0.19). Cochranes Q was estimated at 105 (p < 0.001).

#### Income

Results from the dichotomised meta-analysis of the association between income and depression, including data from 28 studies, can be seen in figure 3. In the pooled estimate, individuals on a low income had a 96% increase in odds of having depression compared to individuals on a high income (OR = 1.96, 95% CI = 1.52-2.53). A sensitivity analysis showed no significant differences in the estimates of risk between high- and low-income economy subgroups (df = 1, p = 0.80). The between-study heterogeneity was estimated at I^2^ = 83% (with a variance of *τ*^2^ 0.35). Cochrane’s Q was estimated at 156 (p < 0.001).

**Figure 3:**
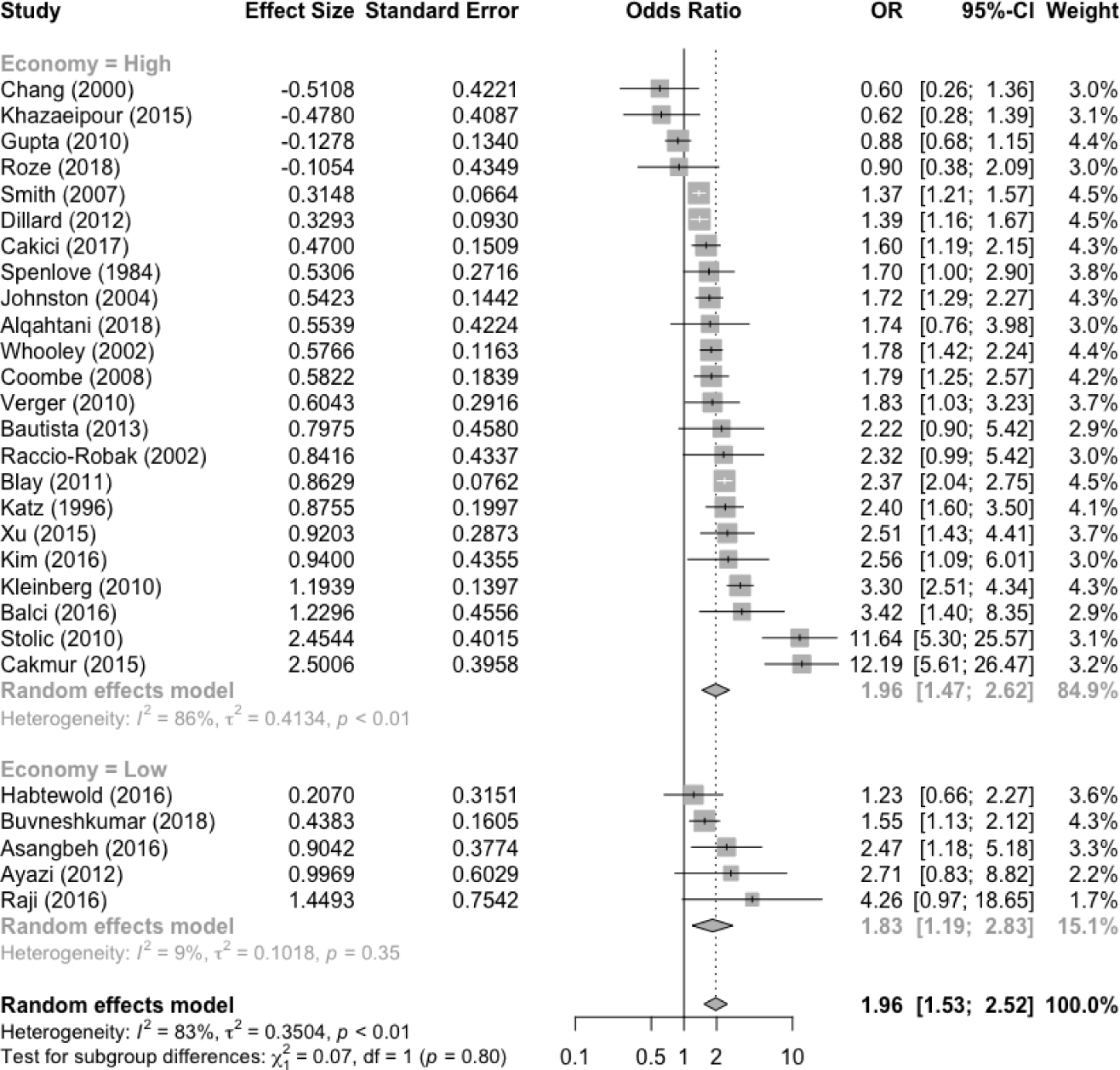
Forrest plot of ORs and CIs of depression risk in people with low vs high income, from dichotomised data. Results are shown for each individual study, a pooled estimate for high- and low-income economy subgroups, and pooled estimate for all included studies.

#### Educational attainment

The results from the dichotomised meta-analyses of association between educational attainment and depression, including 59 studies, can be seen in figure 4.

**Figure 4:**
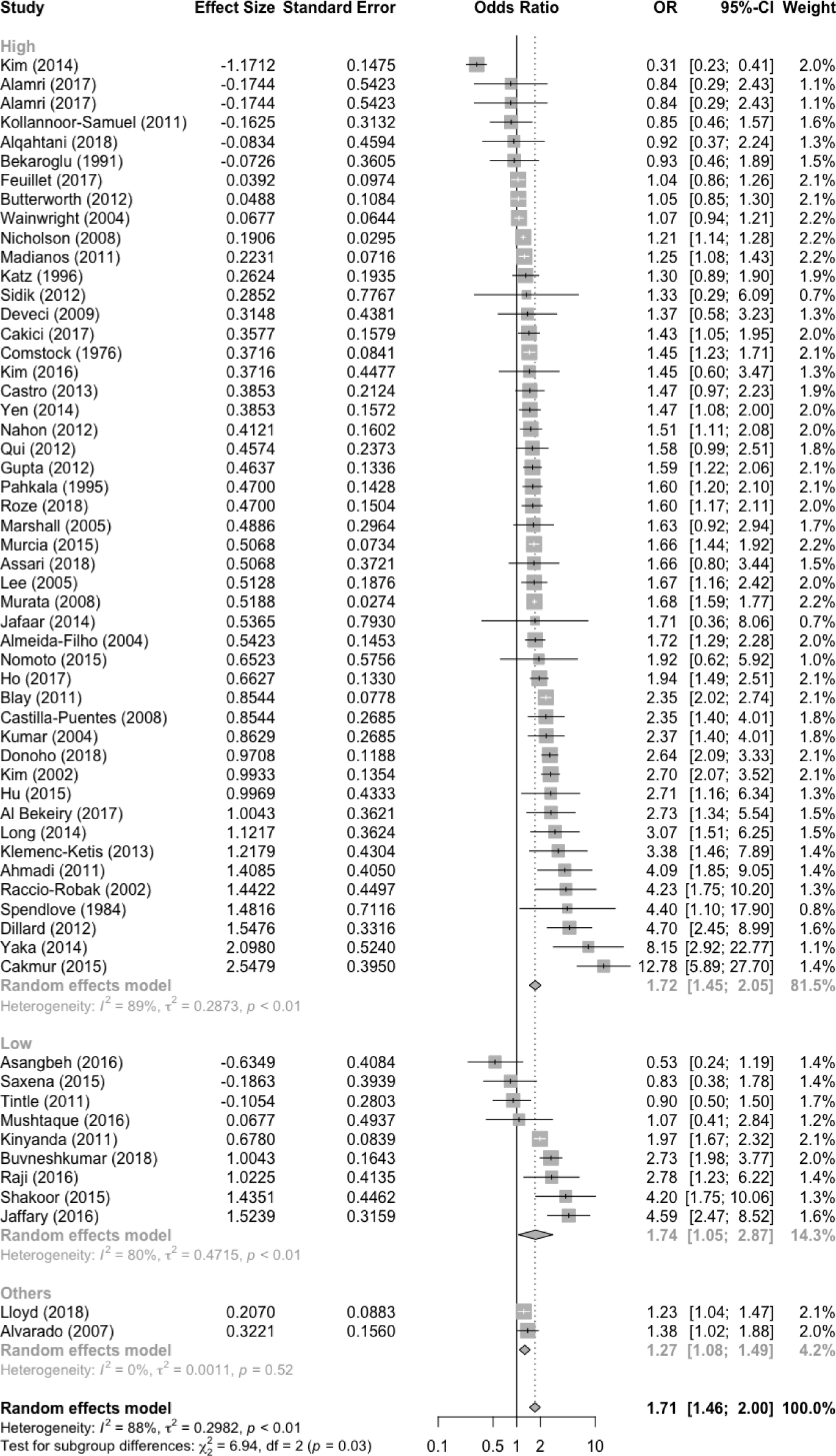
Forrest plot of ORs and CIs of depression risk in people with low educational attainment, from dichotomised data. Results are shown for each individual study, a pooled estimate for high- and low-income economy subgroups, a pooled estimate for studies that could not be placed in either high- and low-income economy subgroup (“others”), and pooled estimate for all included studies.

In the pooled estimate, people with a lower educational attainment had a 69% higher risk of depression compared to people with a higher educational attainment (OR = 1.71, 95% CI = 1.46-2.00). The between-study heterogeneity was estimated at I^2^ = 88% (with a variance of *τ*^2^ 0.29). Cochrane’s Q was estimated at 465 (p < 0.001). Note that two included studies reported education-depression associations in multiple countries, including both high- and low-income economy countries. When these studies were included as a separate subgroup, there was a significant difference between risk estimates of the three economy subgroups (df = 3, p = 0.03), with the pooled estimate for this third subgroup demonstrating relatively weak associations between low educational attainment and increased depression risk. However, without the third subgroup, there was no significant difference between risk estimates of the high- and low-income economy subgroups (df = 1, p = 0.90).

#### Employment

The dichotomised meta-analysis on the association between employment (or unemployment) and depression, including results from 29 studies, can be seen in figure 5.

**Figure 5:**
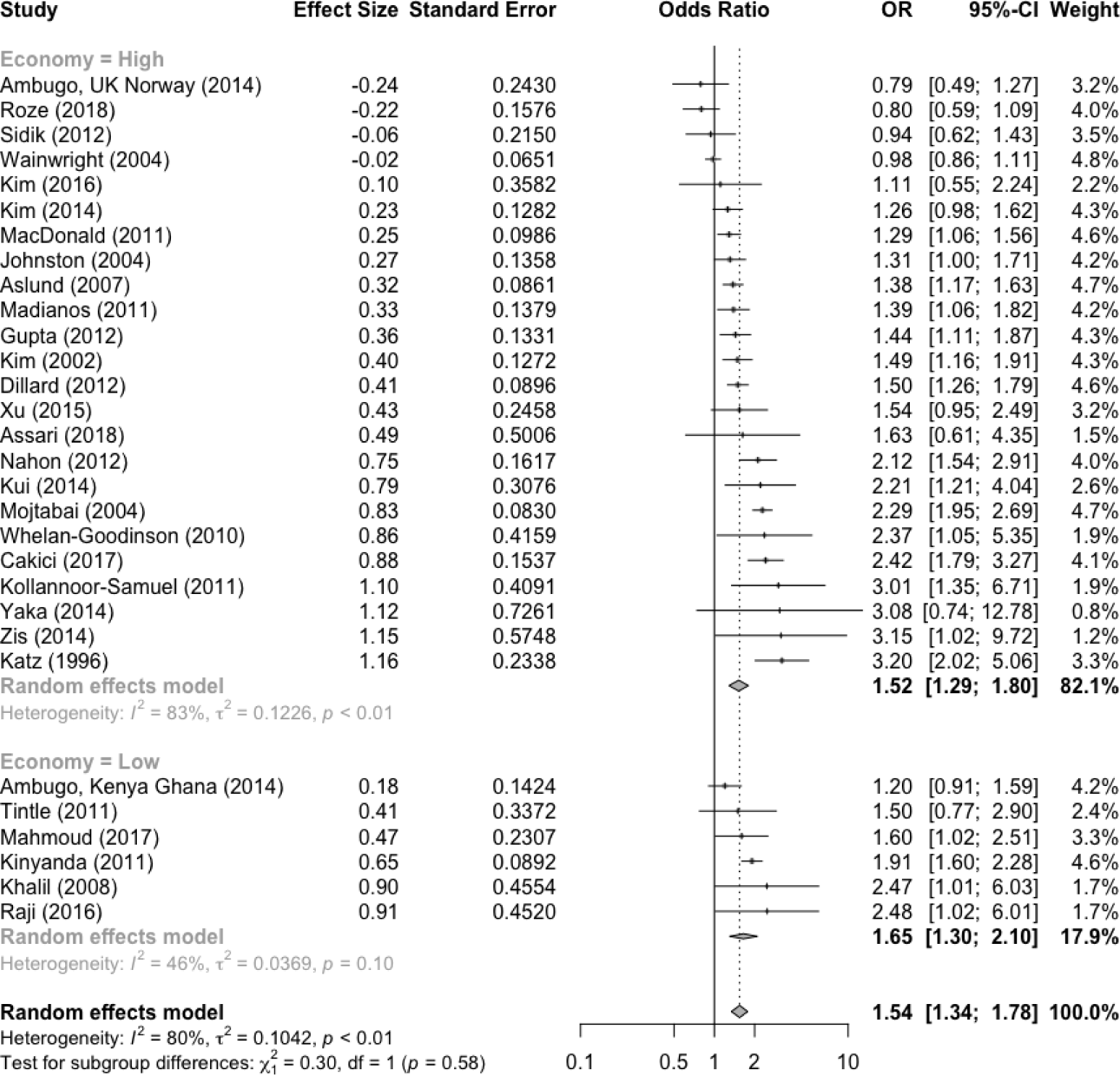
Forrest plot of ORs and CIs of depression risk in people with unemployment, from dichotomised data. Results are shown for each individual study, a pooled estimate for high- and low-income economy subgroups, and pooled estimate for all included studies.

The pooled estimate indicated that people who were unemployed had a 57% higher risk of experiencing depression. The between study heterogeneity was estimated at I^2^ = 81% (with a variance of *τ*^2^ = 0.09). Cochranes Q was estimated at 138 (p < 0.001). There was no significant difference between the risk estimates for low-income economies and high-income economies (df = 1, p = 0.58).

Forest plots of the ordinal and continuous meta-analyses can be found in Additional file 3, tables of narrative and longitudinal narrative reviews, can be found in Additional file 4.

## Discussion

In all but one of the nine separate meta-analyses a consistent association was found between low SES and increased risk of depression. These results were synthesised from 138 studies spanning almost five decades of research, 47 countries across six continents, covering the whole human life span, and including 34 different comorbid diseases.

### Main findings

Low composite SES, income, and educational attainment were all significantly associated with an increased risk of depression in 8 out of 9 meta-analyses. Unemployment was also associated with higher risk of depression. Of the 19 eligible longitudinal studies identified, 15 reported a significant relationship between low SES and higher risk of depression and 7 studies reported a null association. The majority of null associations were reported between educational attainment and depression risk. None of the studies reported a significant relationship between high SES and an increased risk of depression. Overall, the findings from the longitudinal studies and the cross-sectional studies are congruent; most studies report a significant association between low SES and increased risk of depression. Among all 162 reviewed studies, only three (Kim et al. 2014; Kui et al. 2014; Bekaroglu et al. 1991) found a reverse relationship, i.e. low SES being associated with a decreased risk of depression.

The meta-analyses of dichotomised data revealed an increased risk of depression associated with low SES in each of the measures (composite SES, income, education, and unemployment). SES is a composite measure made up of components either directly or indirectly related to income, education, and employment. It is perhaps not surprising that each measure of SES showed a similar direction of association with depression, given these measures are related. Income, education, and employment are, to varying degrees, intercorrelated whilst also each accounting for a proportion of unique variance. For example, having a high level of education makes it more likely to obtain a high income, whilst having a high income makes it easier to retrain or pursue a higher level of education. Having any income at all often relies on being employed, although this does differ from country to country depending on the degree of economic support provided. However, these relationships are far from fixed and many societal and individual factors influence the strength of the relationships between components of SES. When partitioning the risk of depression associated with SES into component risk factors it is important to remember that these components are correlated with one another.

In the present systematic review low income showed the largest magnitude of association with depression (OR = 1.96, 1.53-2.52) than other SES measures. The meta-analyses of studies dividing SES into three groups further supported this finding. Furthermore, there was evidence of a dose dependent pattern, with higher depression risk in medium versus high income groups and low versus across medium income groups (see Additional file 3). Sensitivity analysis showed no significant difference in the risk estimates between high-income economies and low-income economies, suggesting a cross-culturally stable association between low relative income and higher depression risk. A recent review demonstrated an increased risk of depression in populations with higher income inequality (Patel et al. 2018) which was not replicated in this review.

The direction of the effect is unclear from the reviewed studies. However, six out of eight longitudinal studies reported an increased risk of depression following a period of low income (two reported no significant association), suggesting the direction of causation may be from low income to increased risk of depression. The reason behind the strength difference in risk estimates between the SES measures, particularly the much stronger association for low income, is also unclear. It is possible that low income, or poverty, is particularly restrictive in terms of direct and indirect health promoting behaviours and particularly impactful for psychological stress. It is also possible that income has a more direct effect on depression risk as it is often the knock-on effect of other SES measures.

This finding could be important for informing policy intervention attempts. Of all the SES measures, income may be the easiest immediate factor to change. Changing someone’s educational attainment or employment status takes a long time and is difficult, involving changes to the structure of the education system and labour market through macro-economic measures. Preparing an individual for higher educational attainment or a more favourable position within an existing labour market may entail removing impediments based on poor health and carer commitments.

An increasingly appealing intervention against poverty (and low SES) is universal basic income (UBI) (Arnold, 2018). At its core UBI means giving a regular amount of money to an individual without any obligations or commitments necessary. Smaller trials of this intervention have had positive effects on health, wellbeing and SES (Enns et al., 2021; Forget, 2011; Koistinen & Perkiö, 2014). Many of these interventions are relatively short-lived, leaving conclusions on the long-term impact vague. However, larger, and longer interventions are underway in Kenya and the United States, lasting up to 12 years.

Studies on the association between SES and depression are often of an observational nature and as such are prone to biases. However, using these observational studies to inform the design of interventional experimental studies could be a methodologically strong approach. Given the present systematic review found the highest risk of depression to be associated with low income, UBI may present a preventative opportunity in an intervention based experimental set-up.

### Strengths and limitations

The main limitation of the present systematic review is the high levels of heterogeneity in the meta-analyses, which may in-turn reflect differences in study populations, methods or other factors (e.g., cohort effects where the association effect size changes over decades). The analyses involving dichotomous data all have heterogeneity estimates (I^2^) between 81% and 89%. Sensitivity analyses were performed on each of the meta-analyses to investigate if differences in countries’ economies could explain the high-level of heterogeneity. None of the meta-analyses differed significantly in risk estimates between low-income economies and high-income economies. The sensitivity analyses of income and unemployment revealed that most of the heterogeneity were in studies from high-income economies. However, for both meta-analyses there were four times as many studies in the high-income economy group which may be behind the difference in between-study heterogeneity.

A good example of why the heterogeneity may not stem from differences in cultures can be found in one of the studies included in this review; Ambugo et al (2014). The study compares SES and depression associations in four countries (Ghana, Kenya, UK, and Norway) spanning low- and high-income economies, but crucially all with a similar design. The risk estimates were similar and a meta-analysis of the effects reveals relatively little heterogeneity (I^2^ = 50.9, Q = 6.11).

The high levels of between study heterogeneity could instead be due to variation in depression diagnosis screening tools, recruitment settings, and differences in covariate inclusion across studies.

Finally, due to the observational nature of the data it is difficult to confidently infer causality even when including longitudinal data. In the case of investigating the association between a complex disease, such as depression, and a complex phenotype such as SES this is even more pronounced.

The strengths of the present systematic review are its cross cultural, cross age-group, and cross morbidity estimates of the association between SES and depression, as well as providing risk estimates modelled on the largest evidence base to date.

Furthermore, the present meta-analyses are unlikely to be hindered by publication bias, and as a consequence less likely to over-estimate the real risk of depression associated with SES. For many of the included studies, the association between SES and depression was not their primary outcome; this may also explain why a number of studies reported null findings.

Finally, the inclusion of all measures of SES, whether composite or single indicator measures, gives an opportunity to compare the risk of depression across subparts of SES. This, in turn, lays the groundwork for future research to pinpoint which part of an individual’s SES confers the highest risk of depression.

### Future work

Future research should strive to elucidate the direction of causality in the SES-depression association. This could be obtained using cross generation longitudinal population cohort studies, such as Avon Longitudinal Study of Parent and Children with data on several generations of participants.

Another tool for examining causal directions is Mendelian Randomisation (MR), which takes advantage of genetic variants as instrumental variables that are associated with the independent variable (e.g., SES) but not directly with the dependent variable (e.g., depression risk). Previous MR studies into the association between low SES and depression found a significant association between the two factors but did not find a robust indication of causal direction (Harrison et al., 2020). Future MR studies would benefit from stronger genetic instrumental variables and should investigate the possibility of a bidirectional relationship between low SES and depression.

A better understanding of genetic risk of depression and the genetic make-up of SES, will also provide the opportunity to tease apart the social or environmental part of SES that confers risk of depression from the genetic part. This information could aid in pinpointing areas of maximum impact for interventions or even help guide research into preventative treatments of depression.

## Data Availability

All data produced are available online at: https://osf.io/dqnp5/?view_only=ca92b1486ab842b4bbf8c20f9ea4e4c2

## Notes

### Competing Interest Statement

The authors have declared no competing interest.

### Funding Statement

This study was funded by The Wellcome Trust

