## Additional File for "Socioeconomic Status and Depression – A Systematic Review"

### Additional files 1: Data collection table

| **Author** | **Year** | **Journal** | **Country name** | **Country: L, LM, HM, H income?** | **Cohort** | **Setting: Community, clinical, register, etc** | **Study design: Cross sectional, longitudinal** | **Sample size.** | **Number of controls** | **Number of cases.** | **Female** | **Age (preferably margin)** | **Groups (case control)** | **Depression diagnostic tool.** | **Comorbidities.** | **SES measure: single indicator, composite (which?)** | **No. of SES groups** | **Statistical estimates for comparisons.** | **Depression** | **SES** | **Income** | **Occupation** | **Education** | **Variables adjusted for.** | **comments** |
| --- | --- | --- | --- | --- | --- | --- | --- | --- | --- | --- | --- | --- | --- | --- | --- | --- | --- | --- | --- | --- | --- | --- | --- | --- | --- |
| Adewuya et al | 2006 |  |  | LM |  | Clinical | Cross sectional | 102 |  | 28 | 42 | Mean (SD) = 63.22 (10.25) | MDD case/control | MINI | Heart failure | Composite (occupation, educational level, monthly income) | 3 | Chi squared |  | Low: All = 41, with MDD = 16, no MDD = 25  Middle: All = 45, with MDD = 10, no MDD = 35  High: All = 16, with MDD = 2, no MDD = 14 | - | - | - | none |  |
| Ahmadi et al. | 2011 |  |  | HM |  | Clinical | Cross sectional | 114 |  | 49 | 0 | Mean = 34.1 | >17 BDI score(+DSM-V) VS <17 BDI score | Beck Depression Inventory (score 17 cutoff) + DSM-V interview | infertility | Single indicator (economic status and education) | 2 | Logistic regression | - | - |  | - | OR = 0.2; CI 0.06–0.5, Wald = 9.1 P < 0.003 <12th grade VS 12th grade and over | not mentioned |  |
| Alamri, Bari, Ali | 2017 |  |  | H |  | Clinical | Cross sectional | 200 | 176 | 24 | 118 | Mean (SD) = 70.2 (8.1) | DSM-IV MDD vs. not depressed | DSM-IV | Hopitalized elderly | Single indicator (income and education) | 3 | unknown | - | - | RAW DATA (number of people): Low income depressed = 14, Low income not depressed = 98. Median income depressed = 6, Median income not depressed = 50. High income depressed = 4, High income not depressed = 28. P = 0.96 | - | RAW DATA (number of people) Education grade 12 or below: depressed = 19, not depressed = 144. P = 0.97 | not mentioned |  |
| Al Bekairy et al. | 2017 |  |  | H |  | Clinical | Cross sectional | 158 | 70 | 85 | 74 | Mean (SD) = 67.2 (12.6) | Depressed vs not depressed | HADS | Diabetes | Single indicator | 3 | Chi squared | - | - | Income, depression present vs. absent: >10,000 SR: absent = 25, present = 13 5,000 – 10,000 SR: absent = 15, present = 12 <5,000 SR: absent: 36, present = 57 p = 0.035 | Job status depression absent, present: Retired: absent = 33, present = 45 Unemployed: absent = 25, present = 38 Employed: absent = 16, present = 1 p = 0.001 | University degree: absent = 10, present = 8 Secondary and high school: absent = 24, present = 9 Primary school: absent = 19, present = 17 Illiterate: absent = 22, present = 49 p = 0.002 | none |  |

### Additional files 2: Reasons for exclusion

| **Reason for exclusion** | **Number of studies** |
| --- | --- |
| Symptom severity rather than clinical depression | 188 |
| no SES MDD comparison | 78 |
| insufficient reporting of results | 60 |
| Several psychiatric disorders in the experimental group only | 17 |
| Studies not available in English | 10 |
| Studies could not be found. | 4 |
| Data already been published in other included studies | 4 |
| No comparison with a control group | 4 |
| Perinatal depression | 4 |
| Book chapters, comments, or conference proceedings | 4 |
| Reviews | 3 |
| Duplications | 2 |
| Matched cases and controls for SES | 1 |
| Qualitative study | 1 |
| Reported data and results did not match | 1 |

### Additional files 3: Forest plots

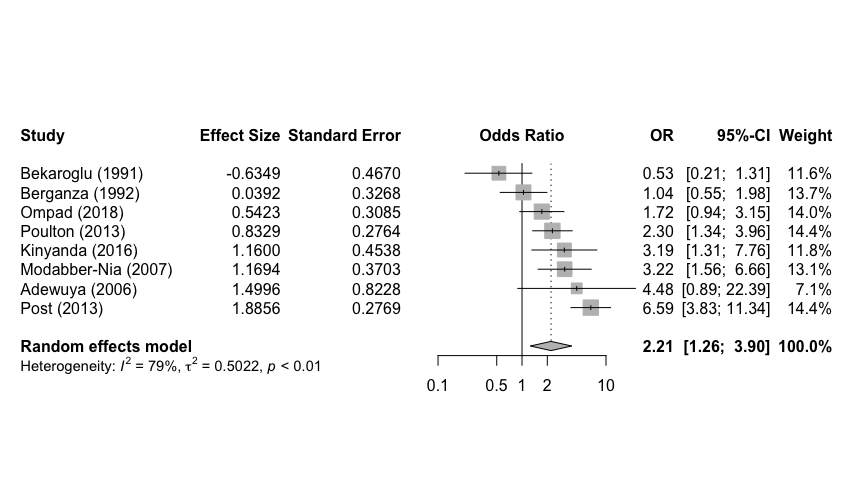

Figure A.3.1: Forest plot of depression risk associated with low versus high SES.

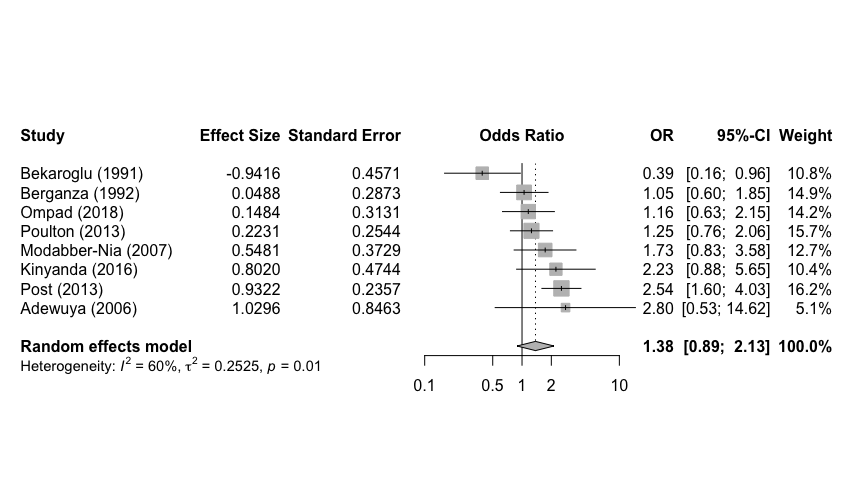

Figure A.3.2: Forest plot of depression risk associated with medium versus high SES.

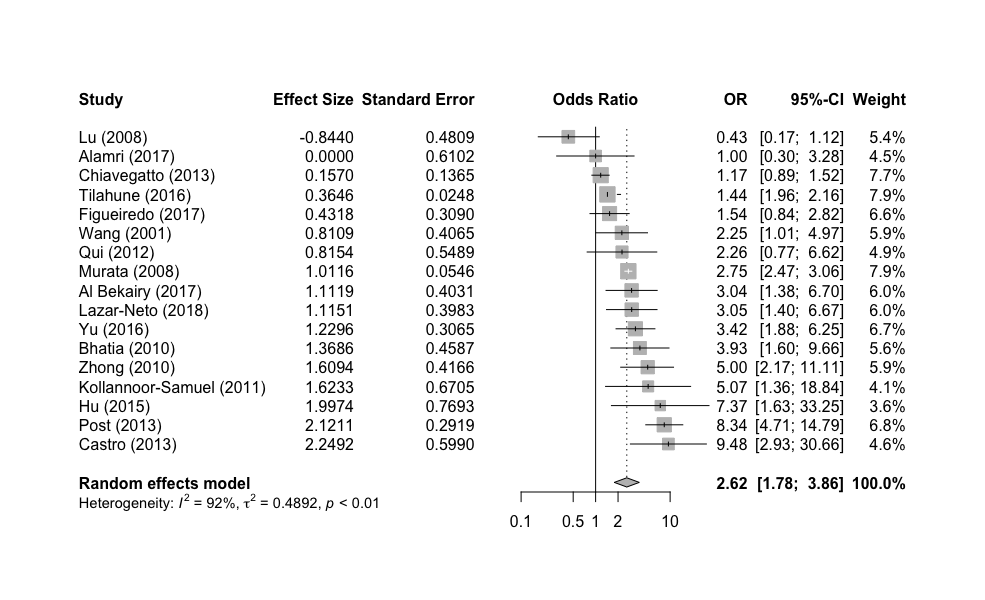

Figure A.3.3: Forest plot of depression risk associated with low versus high income.

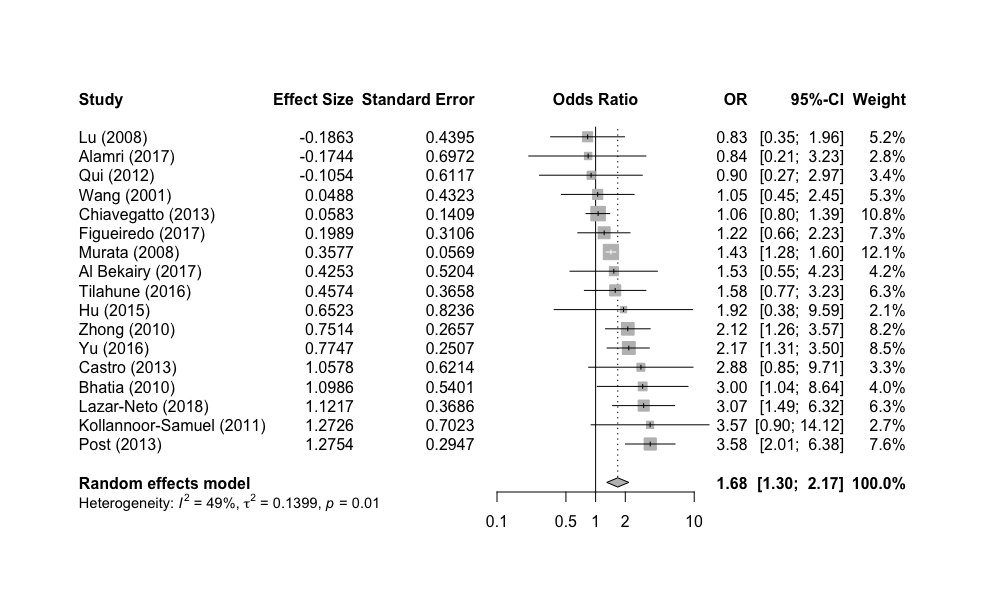

Figure A.3.4: Forest plot of depression risk associated with medium versus high income.

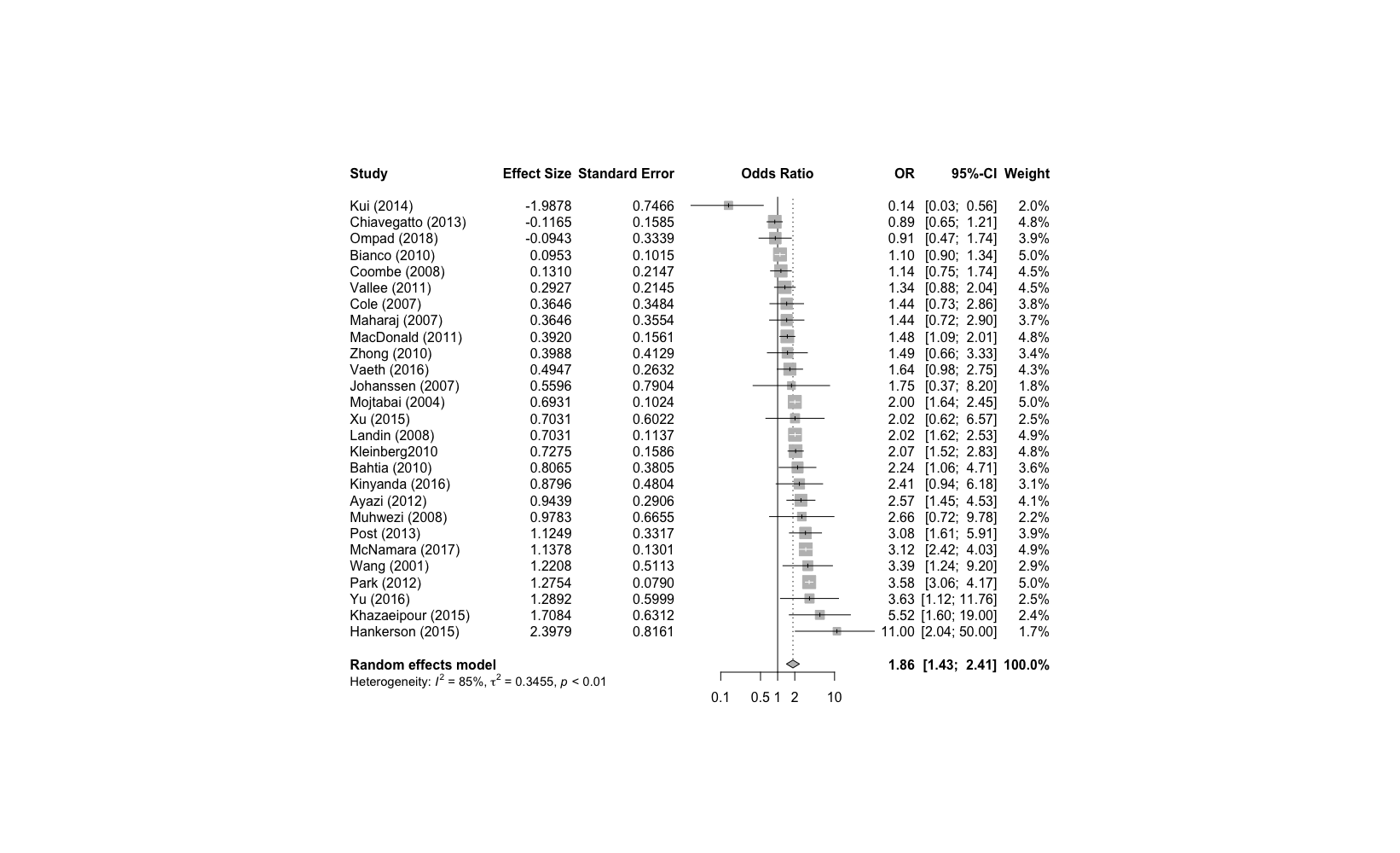

Figure A.3.5: Forest plot of depression risk associated with low versus high educational attainment.

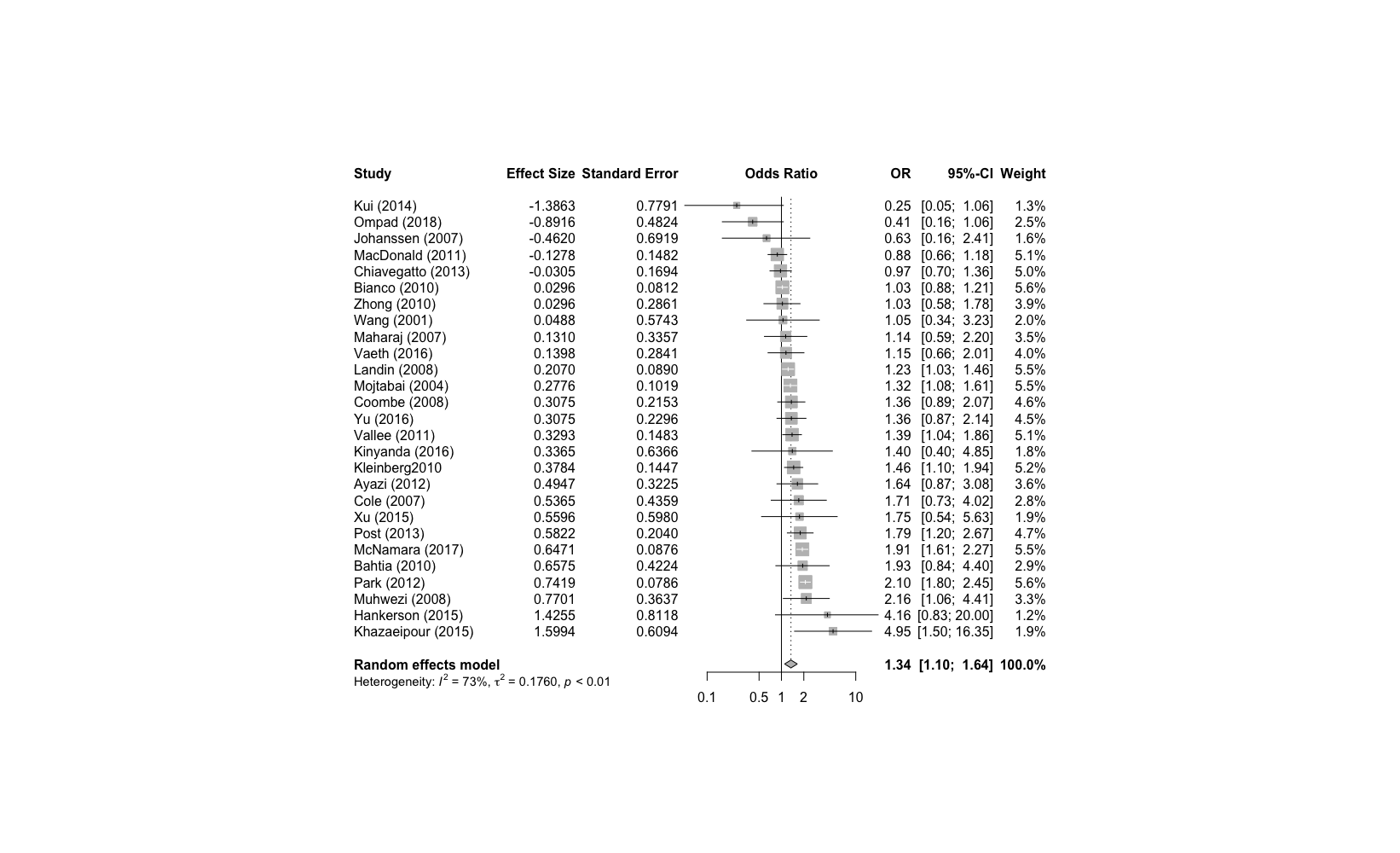

Figure A.3.6: Forest plot of depression risk associated with medium versus high educational attainment.

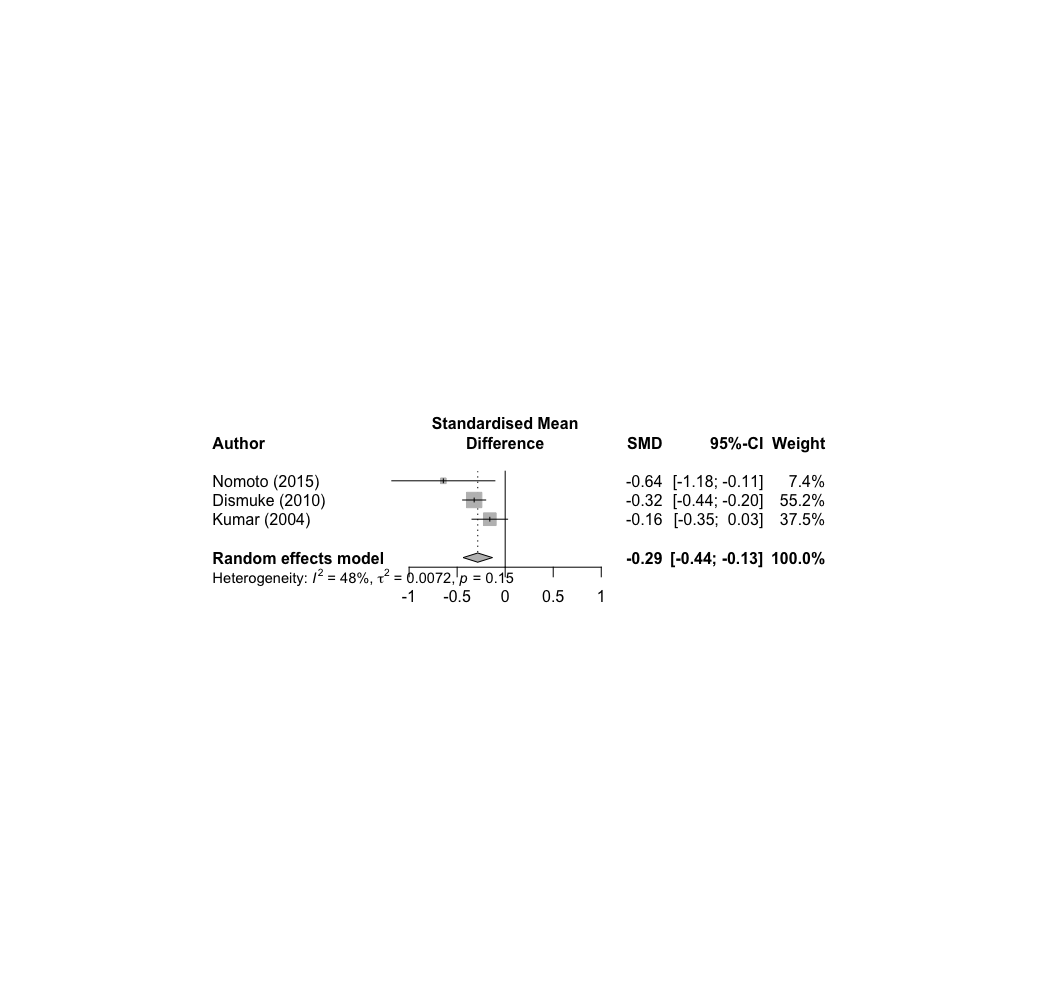

Figure A.3.7: Forest plot of depression risk associated with income based on continuous data.

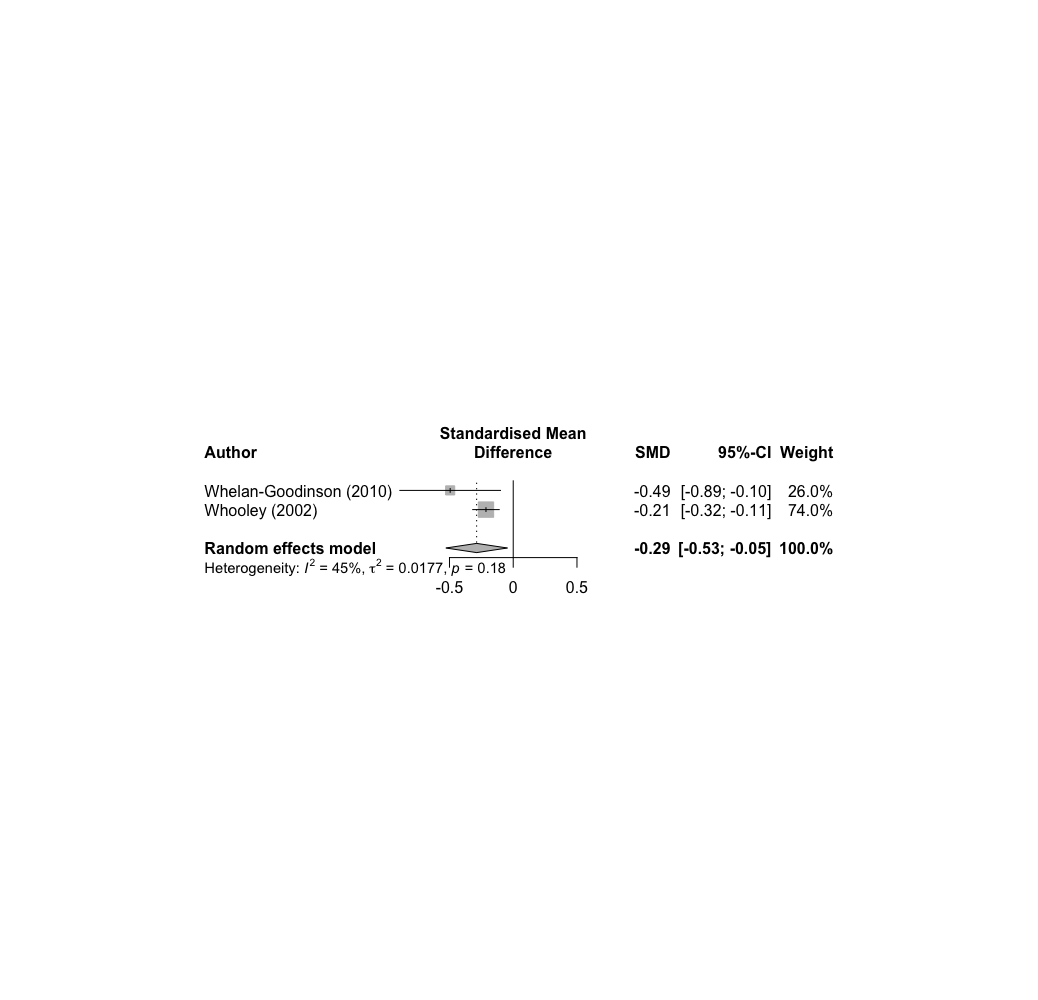

Figure A.3.8: Forest plot of depression risk associated with educational attainment based on continuous data.

### Additional files 4: Longitudinal and narrative review tables

**Longitudinal Narrative Review**

| **Author** | **Country** | **Recruitment setting** | **Years of follow-up** | **Sample size** | **N. of controls** | **N. of cases** | **Result** |
| --- | --- | --- | --- | --- | --- | --- | --- |
| Lee (2016) | Taiwan | Community | 10 | 1,743,948 | 1,727,693 | 16,255 | Income:  Percentage of low-income individuals with MDD (1.73%) significantly higher than non-low-income individuals (0.92%), p < 0.0001.  No significant change in incidence rates across follow-up. |
| Andersen et al. (2011) | Brazil | Community | 6 |  | T1: 2% T2: 4.9% |  | Percent depressed per employment group: 2000 Non-manual = 0.7% 2000 Manual = 1.3% 2000 Non-employed = 11.3% 2006 Non-manual = 2.8% 2006 Manual = 4.7% 2006 Non-employed = 16.8% |
| Stansfeld (2008) | United Kingdom | Community | 3 | 9377 | 9238 | 139 | Occupation:  After adjusting for childhood SES women working in manual labour (compared to non-manual) were at increased risk of depression (OR = 2.53, CI = 1.59-4.04).  No significant difference in risk of depression in men across occupation types. |
| Wang (2012) | Canada | Community | 1 | 2752 | 2682 | 70 | Low income (< $60,000) was not significantly associated with increased risk of depression after follow-up (OR = 1.42, CI = 0.80-2.51). Low educational attainments (< university degree) was not significantly associated with increased risk of depression after follow-up (OR = 2.20, CI = 0.98-4.95). |
| Almeida (2012) | Australia | Community | 2 | 18,758 | 18,483 | 275  (40 recurrent) | No post-school education was not significantly associated with baseline depression (OR = 1.1, CI = 0.7-1.7) or  repeated depression at follow-up (OR = 0.7, CI = 0.3-1.8) |
| Joergensen (2016) | Denmark | Clinical | 30 days (early) 2 years (late) | 87,118 | 77,579 | 9,539 | Low education (corrected for age and sex): Early depression HR (CI) = 1.13 (0.99-1.28).  Late depression HR (CI) = 1.08 (1.03–1.13)  Unemployment (corrected for age, sex, education):  Early depression HR (CI) = 1.57 (1.29–1.90)  Late depression HR (CI) = 1.53 (1.43–1.64)  Income (corrected for age, sex, education, employment) was not significantly associated with  early depression or  late depression. |
| McGormick (2018) | USA | Clinical | 1 | 848 | 682 | 166 | Household income OR (95%) incident depression: <$40,000: OR = 2.08 (1.40–3.10) $40,000 to <$80,000: OR = 0.92 (0.64–1.32) ≥$80,000 (reference)  Education, OR (95%) of incident depression: Did not graduate college: OR = 1.60 (1.13–2.26) College graduate (reference) |
| Anderson (2011) | Sweden | Community | 5 | 30,970 | 28,500 | 2,470 | Risk of depression associated with Socioeconomic status:  Highest: ref.  Second highest: 1.04 (0.86 - 1.26)  Middle: 1.14 (0.95 - 1.36)  Second lowest: 1.41 (1.17 - 1.70)  Lowest: 1.53 (1.25 - 1.88) |
| Batterham (2009) | Australia | Community | 4 | 6,605 | 6,334 | 271 | Not in full time employment was associated with increased risk of depression 4 years later (OR = 2.1, X^2^= 8.3, p = .0040) |
| Hamano (2019) | Sweden | Clinical | 7 | 188,907 | 166,893 | 22,014 | Significant association between both educational attainment and income and development of depression in both men and women. |
| Kosidou (2011) | Sweden | Community | 5 | 21,821 | 21,437 | 384 | Significant association between income and depression in both men and women. No association between educational attainment and depression in either men or women. |
| Wirback (2018) | Sweden | Community | 4 | 169,262 | 162,823 | 6,439 | Disposable Household income  High (ref.): Medium high: OR (95%) = 1.2 (1.1–1.2) Medium: OR = 1.3 (1.2–1.4) Medium low: OR = 1.2 (1.1–1.3) Low: OR (95%) = 1.0 (0.9–1.1)  Parental education High (ref.):  Medium: OR = 1.1 (1.1–1.2)  Low: OR = 1.1 (1.0–1.2) |
| Zhao (2018) | China | Community | 1 | 945 | 825 | 130 | Significant association between poor economic situation and depression (OR = 8.294, CI = 3.12-22.02). No significant association between education and depression. |
| Tani (2016) | Japan | Community | 50+ | 10,458 | 9,095 | 1,363 | Childhood SES (at age 15)  High: ref  Middle: ARR = 1.13 (0.96–1.33)  Low ARR = 1.44 (1.23–1.69) |
| Koster (2006) | Netherlands | Community | 9 | 2,593 | 2189 | 404 | Education  High: ref  Medium: HR = 1.30 (0.98–1.72)  Low: HR = 1.52 (1.17–1.99)  Income  High: ref  Medium: HR = 1.44 (1.11–1.85)  Low: HR = 1.48 (1.13–1.94) |
| Lorant (2018) | Belgium | Community | 7 | 54,190 | 50,288 | 3,902 | Change in:  Poverty: OR = 1.12 (0.97-1.30)  Unemployment: OR = 1.06 (0.81-13.0) |
| Ritsher (2018) | USA | Community | 17 | 306 | 207 | 99 | Neither parent educated beyond high school:  OR = 2.50 (1.46-4.28) |
| Wang (2009) | Canada | Community | 6 | 9,589 | - | - | <13 years education:  OR = 1.83 (1.27-2.64) |
| Daskalakis (2002) | Canada | Community | 4 | 631 | 599-604 | 27-32 | Education (grades):  ≥11: Ref.  <11: OR (95% CI) = 2.14 (1.06, 4.35) |

**Narrative Review**

| **Author** | **Country** | **Recruitment setting** | **Study design** | **N. of controls** | **N. of cases** | **Result** |
| --- | --- | --- | --- | --- | --- | --- |
| Turner et al (1999) | USA | Community | Cross sectional | 1239 | 154 | Occupational level Major Professional: MDD % = 4.1 Lesser Professional: MDD % = 7.9 Minor Professional: MDD % = 9.7 Clerical/Sales: MDD % = 14.5 Skilled/Manual: MDD % = 7.2 Semiskilled/Unskilled: MDD % = 18.6 |
| Wilson et al. (1999) | UK | Community | Cross sectional | 1469 | 417 | Townsend score: -1: OR (ref) = 1 0-1: OR (95% CI) = 2.64 (1.33-5.29) 4-5: OR (95% CI) = 1.30 (0.78-2.17) 6-7: OR (95% CI) = 1.28 (0.78-2.11) 8-9: OR (95% CI) = 1.89 (1.16-3.08) 10-13: OR (95% CI) = 2.0 (1.16-3.47) |
| Goodman et al. (2003) | USA | Community | Cross sectional | 13602 | 1510 | Parental income: Depression relative risk (95%CI) Q5: OR = 1 (ref.) Q4: OR = 1.21 (1.36, 1.95) Q3: OR = 1.46 (1.23, 1.74) Q2: OR = 1.63 (1.36, 1.95) Q1: OR = 2.07 (1.73, 2.47)  Parental education OR (95%CI): Professional Degree: OR = 1 (ref.) College Graduate: OR = 1.5 (1.10, 1.88)  > High School, < College: OR = 1.6 (1.31, 1.98)  High School: OR = 1.95 (1.59, 2.40)  < High School: OR = 2.95 (2.37, 3.67) |
| Beard et al. (2008) | USA | Community | Cross sectional |  | 605 | Household income  $100,000+: OR = 1.00 (ref.), $75,000–$99,999: OR (CI) = 1.27 (0.69–2.33), $50,000–$74,999: OR (CI) = 1.67 (0.96–2.88), $40,000–$49,999: OR (CI) = 3.46 (1.92–6.22), $30,000–$39,999: OR (CI) = 4.22 (2.39–7.45), $20,000–$29,999: OR (CI) = 3.39 (1.96–5.86),  <$20,000: OR (CI) = 5.05 (3.04–8.41)  Educational attainment  Graduate work: OR = 1.00 (ref.), Bachelor of arts degree: OR (CI) = 1.46 (0.90–2.39), Some college: OR (CI) = 1.99 (1.20–3.32), High school/GED: OR (CI) = 1.97 (1.19–3.29),  <High school: OR (CI) = 3.22 (1.83–5.69) |
| Pikhartova et al. (2009) | Czechia | Community | Cross sectional | 6123 | 1493 | Deprivation level, OR (95% CI): 0: OR = 1 (ref) 1–2: OR = 1.91 (1.61–2.26) 3–5: OR = 2.76 (2.36–3.23) 6–12: OR = 4.87 (4.07–5.82)  Deprivation, OR (95% CI): Primary or less: OR = 1 (ref) Vocational: OR = 0.73 (0.62–0.86) Secondary: OR = 0.63 (0.53–0.75) University: OR = 0.48 (0.38–0.62) |
| Poongothai et al. (2009) | India | Community | Cross sectional | 21608 | 3847 | Income level, OR (95% CI): > 20000: OR = 1 (ref.) 10001–20000: OR = 1.15 (0.54–2.43) 5000–10000: OR = 1.41 (0.70–2.81) < 5000: OR = 2.44 (1.24–4.81)  Education status, OR (95% CI): Professional: OR = 1 (ref) Post graduate: OR = 1.09 (0.52–2.31) Graduate: OR = 1.35 (0.70–2.59) SSC: OR = 2.00 (1.05–3.82) Below SSC: OR = 2.66 (1.40–5.07) Illiterate: OR = 4.77 (2.50–9.11) |
| Braich et al. (2011) | India | Community |  |  |  | Income: <2500: OR = 1 (ref.) 2500-6000: OR = 0.31 6000-15000: OR = 0.17 15000-24000: OR = 0.06 >24000: OR = 0.68 |
| Andersen et al. (2011) | Brazil | Community | Longitudinal (6yrs) |  | T1: 2% T2: 4.9% | Percent depressed per employment group: 2000 Non-manual = 0.7% 2000 Manual = 1.3% 2000 Non-employed = 11.3% 2006 Non-manual = 2.8% 2006 Manual = 4.7% 2006 Non-employed = 16.8% |
| Yunming et al. (2012) | China | Community | Cross sectional | 1085 | 401 | Max income (RMB) 0–300: OR = 1(ref) 301–500 : OR = 1.11 (0.21–5.79) 501–1000 : OR = 1.12 (0.26–4.90) 1001–1500: OR = 0.73 (0.20–2.66) 1500 or above: OR = 0.42 (0.15–1.16)  Education level  No education: OR = 1 (ref) Elementary school: OR = 0.96 (0.13–7.27) Primary school: OR = 1.34 (0.31–5.71) Secondary school: OR = 1.40 (0.27–7.23) Post-secondary school/above: OR = 0.85 (0.28–2.62) |
| Alonso-Moran et al. (2014) |  | Clinical | Cross sectional |  | 12392 | Deprivation index (95% CI) 1: OR = 1 2: OR = 0.95 (0.88, 1.03) 3: OR = 1.01 (0.93, 1.09) 4: OR = 0.93 (0.86, 1.01) 5: OR = 0.96 (0.89, 1.03) |
| Shi et al. (2015) | China | Clinical | Cross sectional | 3639 | 3800 | Social status (%) Semi skilled and unskilled workers: MDD = 9.21,Control = 11.31 Skilled manual employees: MDD = 28.52, Control = 26.2 Administrative personnel, minor profesionals, clerical and sales workers: MDD = 17.24, Control = 14.1 Executives, business owners, major profesionnals: MDD = 27.45, Control = 29.07 Other: MDD = 17.57, control = 19.31  Education (%): no education or pre-school: MDD = 6.82, Control = 3.97 primary school or below: MDD = 18.05, Control = 12.95 junior middle school: MDD = 31.27, Control = 32.92 senior middle school : MDD = 29.24, Control = 30.26 junior college: MDD = 8.85, Control = 11 bachelor degree: MDD = 4.92, Control = 7.89  master degree or above: MDD = 0.85, Control = 1 |
| Magklara et al. (2015) | Greece | Community | Cross sectional | 2181 | 246 | Father’s employment status OR (95%): Public sector employee: 1.00 (ref.) Private sector employee: 1.29 (0.85-1.95) Self-employed: 0.92 (0.62-1.37) Retired: 0.41 (0.17-1.00) Unemployed/ Other:1.02 (0.58-1.80)  Father’s educational level OR (95% CI): Primary: 1.00 (ref) Secondary Basic: 1.03 (0.56-1.85) Secondary Complete: 1.01 (0.63-1.62) Technological degree: 1.41 (0.81-2.45) University degree: 0.94 (0.57-1.54) |
| Freeman et al. (2006) | Spain Finland Poland | Community | Cross sectional | Finland = 1854 Poland = 3726 Spain = 4073 | Finland = 80 Poland = 214 Spain = 510 | SES (0–110) index Finland: OR (95% CI) = 0.991 (0.98–0.997)  SES (0–110) index Poland: OR (95% CI) = 0.986 (0.975–0.99)  SES (0–110) index Spain: OR (95% CI) = 0.989 (0.984–0.995)  Income Quintile Finland: OR (95% CI) = 0.84 (0.714–0.987)  Income Quintile Poland: OR (95% CI) = 0.84 (0.71–0.98)  Income Quintile Spain: OR (95% CI) = 1.0 (0.914–1.09)  Yrs of Education Finland: OR (95% CI) = 0.94 (0.89–0.985)  Yrs of Education Poland: OR (95% CI) = 0.934 (0.88–0.983)  Yrs of Education Spain: OR (95% CI) = 0.913 (0.887–0.939) |
| Altino et al. (2017) |  | Clinical | Cross sectional | 97 | 23 | Mean number of years of education: Depressed: 6 (4–8) Not-depressed: 11 (4–15) p = 0.011 |
| Lotfaliany et al. (2018) |  | Community | Cross sectional | 39302 | 2508 | Wealth quantile, OR (95% CI) Q1: OR = 1.57 (1.33,1.84) Q2 : OR = 1.50 (1.28,1.76)  Q3 : OR = 1.43 (1.22,1.67) Q4 : OR = 1.16 (0.99,1.36) Q5: OR = 1 (Ref.)  Education, OR (95% CI) No formal education: OR = 1.71 (1.31,2.22) Less than primary level: OR = 1.68 (1.28,2.21) Primary level: OR = 1.33 (1.01,1.74) Secondary level: OR = 1.33 (1.04,1.71) Tertiary level: OR = 1 (Ref.) |
| Averina et al. (2005) | Russia | Community | Cross sectional | 2907 | 798 | Education:  Secondary: ref.  Secondary professional: OR = 0.8 (0.7–1.0)  High: OR = 0.9 (0.7–1.1)  Income:  High salary: ref.  Average salary: OR = 1.5 (1.02–2.2)  Low salary: OR = 1.5 (1.04–2.0)  Retirement pension: OR = 1.5 (1.02–2.3)  Other (unknown income): OR = 1.3 (0.9–2.0) |
| Fernandez-Pujals (2015) | UK | Community | Cross sectional | 17,472 | 2,726 | INCOME, OR (95% CI)  >10k = 1.28 (0.96–1.66)  10k-30k = 1.00 (ref)  30k-50k = 0.78 (0.64–0.94)  50k-70k = 0.69 (0.54–0.85)  < 70k = 0.64 (0.47–0.81)  EDUCATION, OR (95% CI)  University Degree = 1.00 (ref)  Post-Secondary = 1.03 (0.85–1.22)  Upper Secondary = 0.68 (0.53–0.86)  Lower Secondary = 0.74 (0.57–0.93)  Primary = 0.65 (0.48–0.83)  SCOTTISH INDEX OF MULTIPLE DEPRIVATION OR (95% CI)  Most deprived quintile = 1.17 (0.88–1.46)  2nd Most deprived quintile = 1.03 (0.80–1.28)  Median quintile = 1.00 (ref)  2nd Least deprived quintile = 0.79 (0.62–0.97)  Least deprived quintile = 0.92 (0.73–1.12) |
| Fillenbaum (2013) | Brazil | Community | Cross sectional | 3549 | 2,269 | OLS regression of depression and continuous income:  F = 193.4, p < 0.0001 |
| Ediz (2017) | Turkey | Community | Cross sectional | 713 | 215 | Good economic situation: ref.  Moderate economic situation: OR (95% CI) = 1.4 (1.008–1.898)  Poor economic situation: OR (95% CI) = 2.9 (1.438–5.683) |
| Gooseby (2013) | USA | Community | Cross sectional | Total sample = 4,339 | - | Childhood SES, parental education:  Relative risk ratio = 0.98, p > 0.1 |
| Liu (2015) | China | Community | Cross sectional | 15,856 | 176 | EDUCATION  Illiterate: ref.  Primary school: OR (95% CI) = 0.655 (0.383–1.119)  Secondary school: OR (95% CI) = 0.393 (0.240–0.645)  High school: OR (95% CI) = 0.349 (0.210–0.581)  College and above: OR (95% CI) = 0.171 (0.099–0.294)  PERSONAL MONTHLY INCOME (RMBS)  0–1499: ref.  1500–2999: OR (95% CI) = 0.750 (0.544–1.035)  3000–4999: OR (95% CI) = 0.337 (0.199–0.570)  5000–7999: OR (95% CI) = 0.448 (0.195–1.030)  8000 and over: OR (95% CI) = 0.553 (0.135–2.264) |
| Hudson (2010) | USA | Community | Cross sectional | Total sample = 2268 | - | Parental Education (Years): OR (95% CI) = 1.08 (1.02- 1.14)  Household Income (log): OR (95% CI) = 0.69 (0.52- 0.94)  Education (Years): OR (95% CI) = 1.00 (0.95- 1.05) |
